# Retrospective Clinical Evaluation of Four Lateral Flow Assays for the Detection of SARS-CoV-2 Antibodies

**DOI:** 10.1101/2020.07.01.20129882

**Authors:** Kathrine McAulay, Andrew Bryan, Alexander L. Greninger, Francisca Grill, Douglas Lake, Erin J. Kaleta, Thomas E. Grys

**Affiliations:** Department of Laboratory Medicine and Pathology, Mayo Clinic, Phoenix, AZ, USA; Department of Laboratory Medicine, University of Washington School of Medicine, Seattle, WA, USA; Vaccine and Infectious Disease Division, Fred Hutchison Cancer Research Center, Seattle, WA, USA; School of Life Sciences, Arizona State University, Tempe, AZ, USA

**Keywords:** SARS-CoV-2, COVID-19, lateral flow, immunoassay, serology, IgG

## Abstract

Coronavirus disease 2019 (COVID-19) is a potentially life-threatening respiratory infection caused by severe acute respiratory coronavirus 2 (SARS-CoV-2), for which numerous serologic assays are available. In a CLIA laboratory setting, we used a retrospective sample set (n = 457) to evaluate two lateral flow immunoassays (LFIAs; two iterations of Rapid Response™ COVID-19 Test Cassette, BTNX Inc.) and a subset of to evaluate SARS-COV-2 IgG/IgM Rapid Test, ACON Laboratories (n = 200); and Standard Q COVID-19 IgM/IgG Duo, SD BIOSENSOR (n = 155) for their capacity to detect of SARS-CoV-2 IgG. In a cohort of primarily hospitalized patients with RT-PCR confirmed COVID-19, the BTNX assays demonstrated 95% and 92% agreement with the Abbott SARS-CoV-2 IgG assay and sensitivity was highest at ≥ 14 days from symptom onset [BTNX kit 1, 95%; BTNX kit 2, 91%]. ACON and SD assays demonstrated 99% and 100% agreement with the Abbott assay at ≥ 14 days from symptom onset. Specificity was measured using 74 specimens collected prior to SARS-CoV-2 circulation in the United States and 31 “cross-reactivity challenge” specimens, including those from patients with a history of seasonal coronavirus infection and was 98% for BTNX kit 1 and ACON and 100% for BTNX kit 2 and SD. Taken with data from EUA assays, these results suggest that LFIAs may provide adequate results for rapid detection of SARS-CoV-2. Replicating these results in fingerstick blood in outpatient populations, would further support the possibility that LFIAs may be useful to increase access to serologic testing

## 1. Introduction

Severe acute respiratory syndrome coronavirus 2 (SARS-CoV-2) emerged in 2019 as the causative agent of coronavirus disease 2019 (COVID-19), a pandemic respiratory infection resulting in over 10 million cases and 500,000 deaths between November 2019 and June 2020 (WHO, 2020).

Antibody detection is currently being implemented in many clinical centers to aid in identification of recent disease and to investigate population seroprevalence.

Accurate laboratory tests impact clinical decision-making, and understanding performance of a test is essential to determination of when to use the test and what the results might mean. For example, specificity is of particular importance in a low prevalence setting (Farnsworth and Anderson, 2020). Lateral flow immunoassays (LFIAs) are an attractive alternative or supplement to automated ELISA and chemiluminescence assays as they require less operator skill and for their potential utility in a point of care (POC) setting. Here we evaluated four LFIAs for the detection of anti-SARS-CoV-2 IgG in clinical samples.

## 2. Materials and Methods

### 2.1. Patient population and clinical specimens

De-identified, presumptive positive specimens (n = 352) from 62 individuals with RT-PCR-confirmed COVID-19 were kindly shared by the Department of Laboratory Medicine at the University of Washington School of Medicine (Seattle, WA) with limited metadata, such as Abbott SARS-CoV-2 IgG immunoassay results and the number of days since symptom onset. These consisted of 250 plasma, 77 serum, and 21 whole blood specimens (a further four unknown specimens were assumed to be either serum or plasma), were received frozen, and underwent either one or two freeze-thaw cycles prior to testing. Specificity specimens were obtained from two sources: 74 excess clinical serum specimens collected and stored in 2018, and 31 “cross-reactivity challenge” specimens collected between March and April 2020. Among these 105 specimens, there were 27 from individuals with a history of seasonal coronavirus infection (as determined by a syndromic respiratory PCR test) within 3 years prior to collection (HKU1, n = 13; NL63, n = 6; OC43, n = 6; 229E, n = 2), and 4 specimens reactive for rheumatoid factor, HIV-1 antibody, HAV total antibody, HBV core total antibody and surface antibody, HCV antibody and/or HSV2 antibody. These specimens were tested after 0, 1, or 2 freeze thaw cycles.

### 2.2. Lateral Flow Immunoassays (LFIAs)

Rapid Response™ COVID-19 Test Cassette (BTNX Inc.): We tested two different iterations of this kit, hereafter referred to as BTNX kit 1 and BTNX kit 2. LFIAs were performed according to the manufacturer’s instructions. Briefly, for BTNx kit 1, 10 µL serum, plasma, or whole blood was transferred to the sample well, followed by one drop of assay buffer; results were read and interpreted after 10-15 min. For BTNx kit 2, 5 µL serum, plasma, or whole blood was transferred to the sample well, followed by two drops of assay buffer; results were read after 15 min.

SARS-COV-2 IgG/IgM Rapid Test (ACON Laboratories): For this assay, hereafter referred to as ACON, 10 µL serum or plasma, or 15 µL whole blood was transferred to the specimen well and then two drops of buffer were added to the buffer well; results were read after 10-15 min.

Standard Q COVID-19 IgM/IgG Duo (SD BIOSENSOR): This kit is supplied as individual IgM and IgG cartridges; only the IgG cartridges were evaluated in this study and are hereafter referred to as SD. For this assay, 10 µL serum, plasma, or whole blood was transferred to the specimen well and then two to three drops of buffer were added to the buffer well; results were read and interpreted after 10-15 minutes.

Results for all assays were interpreted by two readers (KM, TG) and photographed for reference, with the exception of the SD assay, for which the first 30 assays were interpreted by one reader only and not photographed. Both readers were essentially blinded, in that the sample metadata (time to disease onset and Abbott results) were not revealed until reading was complete. Images of discrepant specimens were read by a third independent and blinded individual (FG) and the consensus between two readers was recorded as the final result. A representative image of a positive result on all four assays is shown in Figure 1.

**Figure 1.**
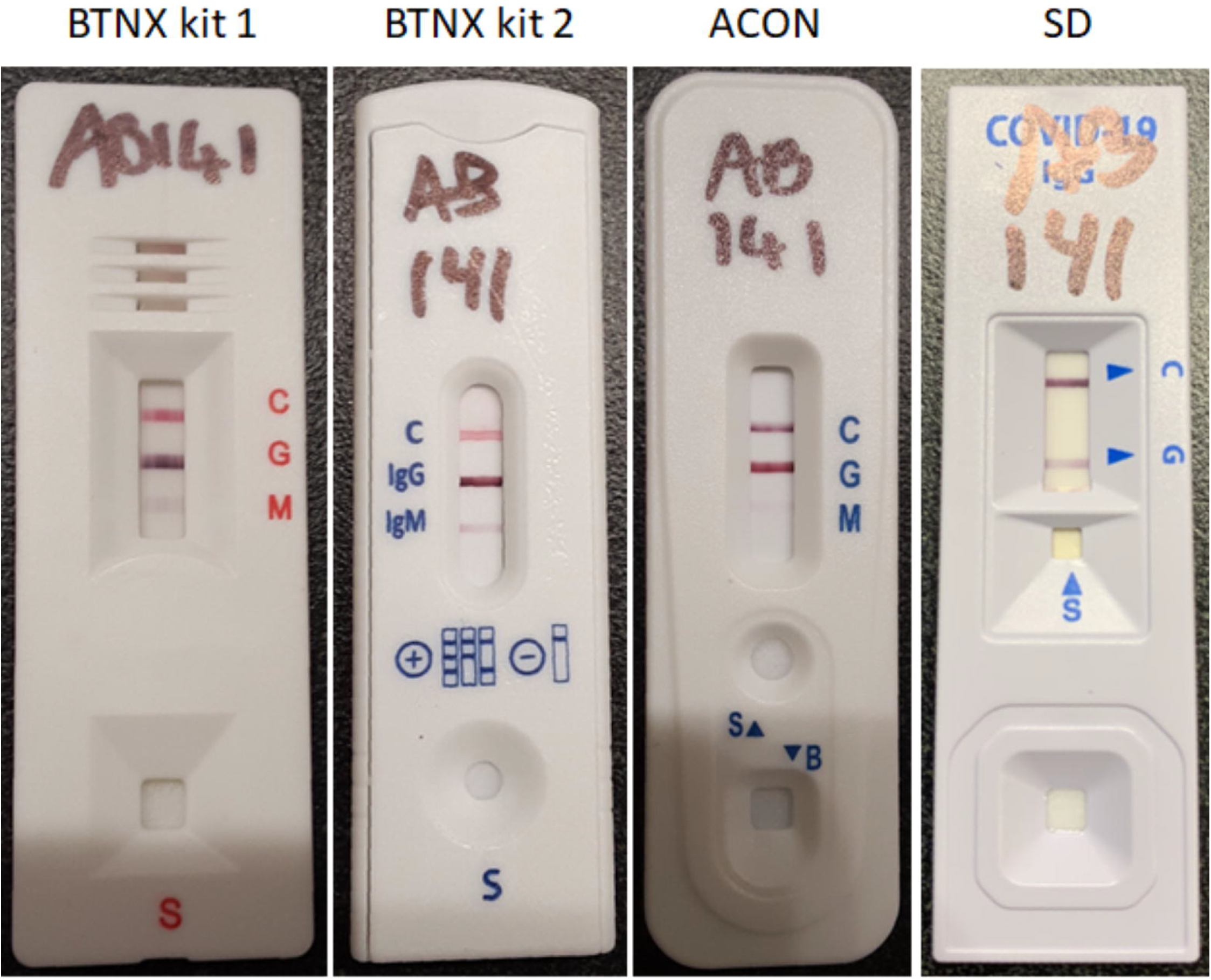
A plasma sample from an RT-PCR confirmed COVID-19 case, collected 14 days after symptom onset and tested on (left to right) Rapid Response™ COVID-19 Test Cassette (BTNX Inc.) Kit 1, Rapid Response™ COVID-19 Test Cassette (BTNX Inc.) Kit 2, SARS-COV-2 IgG/IgM Rapid Test (ACON Laboratories), and Standard Q COVID-19 IgM/IgG Duo (SD BIOSENSOR)

### 2.3. IgM detection

All assays tested also offered IgM detection. The SD IgM cartridge was not evaluated here, but ACON and both BTNX kits included IgM in the same cartridge; however, as IgM results were variable across all assays, we opted to focus on IgG for the purpose of this evaluation.

## 3. Results

### 3.1. BTNX Sensitivity

Sensitivity of the BTNX assays was evaluated using specimens from a primarily hospitalized cohort of individuals with RT-PCR confirmed COVID-19 (Seattle cohort, n = 352) and stratified by the number of days since symptom onset. Sensitivity of BTNX kit 1 at <7 days since symptom onset (n = 154) was 16% (95% CI: 10-22%), at 7-13 days (n = 103) it was 48% (38-58%), and ≥14 (n = 95) days it was 95% (88-98%). Sensitivity of BTNX kit 2 at the same time points was 13% (8-19%), 50% (40-60%), and 91% (83-96%), respectively.

We then compared assay performance to that of the Abbott SARS-CoV-2 IgG assay, which holds and Emergency Use Authorization (EUA) from the FDA and for which optical density (OD) values and interpretations for 268 of these specimens. For a number of samples, >1 sample result was available from the same patient on the same day since symptom onset. When this occurred, the mean OD value was determined and assigned to all samples collected that day. We reviewed the sample specific OD and mean OD for 157 specimens for which both values were available and found that taking the mean did not alter the interpretation in any case; therefore, we opted to use the mean data for comparison with LFIAs. Overall agreement with the Abbott assay was 95% (Cohen’s Kappa, 0.90 [95% CI: 0.85-0.96]) for BTNX kit 1 and 92% (Cohen’s Kappa, 0.84 [0.77-0.90]) for BTNX kit 2.

### 3.2. Sensitivity at ≥14 days since symptom onset

Based on our observation that BTNX kit performance was substantially better for specimens collected ≥14 days after symptom onset and to focus on a sample set in which most patients would be expected to have seroconverted, sensitivity was subsequently addressed for the remaining kits only on specimens collected ≥14 days after symptom onset (n = 95, only 50 of these were tested using the SD assay). This amounted to 95% (88-98%) and 92% (81-98%) sensitivity for ACON and SD, respectively. LFIA results are summarized in Table 1 and listed in full in supplementary table S1.

**Table 1.**
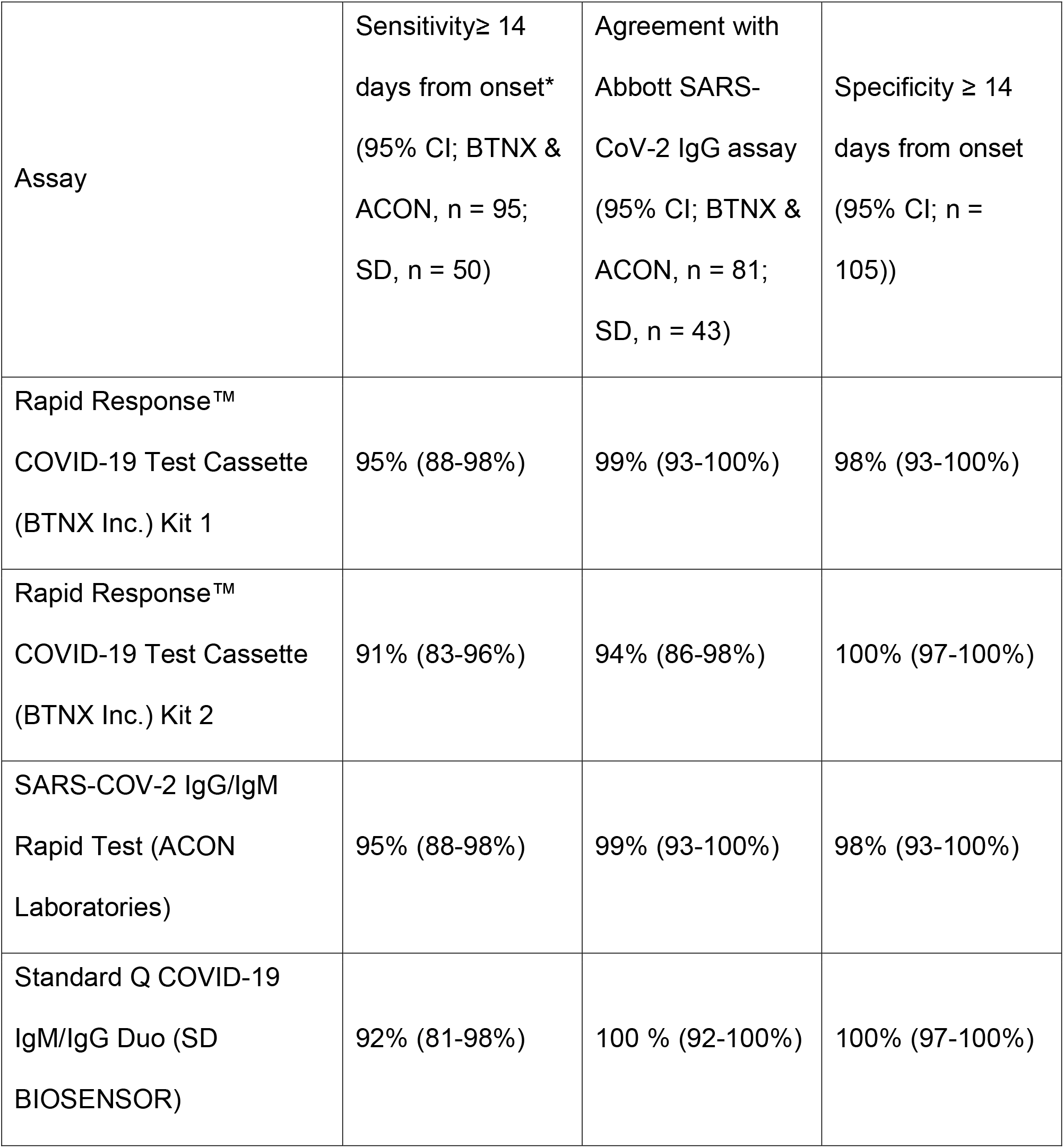
Summarized performance characteristics of four SARS-CoV-2 IgG lateral flow immunoassays. *Where a “true positive” is a specimen collected ≥ 14 days after symptom onset from an RT-PCR confirmed COVID-19 case

| Assay | Sensitivity ≥ 14 days from onset* (95% CI; BTNX & ACON, n = 95; SD, n = 50) | Agreement with Abbott SARS-CoV-2 IgG assay (95% CI; BTNX & ACON, n = 81; SD, n = 43) | Specificity ≥ 14 days from onset (95% CI; n = 105)) |
| --- | --- | --- | --- |
| Rapid Response™ COVID-19 Test Cassette (BTNX Inc.) Kit 1 | 95% (88-98%) | 99% (93-100%) | 98% (93-100%) |
| Rapid Response™ COVID-19 Test Cassette (BTNX Inc.) Kit 2 | 91% (83-96%) | 94% (86-98%) | 100% (97-100%) |
| SARS-COV-2 IgG/IgM Rapid Test (ACON Laboratories) | 95% (88-98%) | 99% (93-100%) | 98% (93-100%) |
| Standard Q COVID-19 IgM/IgG Duo (SD BIOSENSOR) | 92% (81-98%) | 100 % (92-100%) | 100% (97-100%) |

Abbott results were available for 83 of the 95 specimens collected ≥14 days after symptom onset (43 of the 50 tested by SD) and agreement was as follows: BTNX kit 1, 99% (93-100%); BTNX kit 2, 94% (86-98%); ACON, 99% (93-100%); and SD, 100% (92-100%).

### 3.3. Specificity

As all samples in the Seattle cohort were from laboratory or clinically confirmed COVID-19 cases, a different set of specimens was used to assess specificity (n = 105), including 74 collected prior to recognized circulation of SARS-CoV-2 in the United States and 31 “cross-reactivity challenge specimens” from individuals with a history of seasonal coronavirus infection or other potentially cross-reactive antibodies. Two false positive IgG results were observed with BTNX kit 1, amounting to 98% (93-100%) specificity. Of note, one additional specimen generated a pink line (supplementary figure S1), where a purple colored line is the expected result. This was recorded as invalid rather than a positive result, since it was not consistent with the operating parameters of the assay. None of the 31 “cross-reactivity challenge specimens” generated a positive IgG signal in this assay. The ACON assay also generated two false positive results, one of which was from a patient with a recent history of coronavirus HKU1 infection and the other was from a patient whose serum was reactive for HIV-1, HAV, HBV, and HCV antibodies. This assay therefore also achieved 98% (93-100%) specificity. False positive results are summarized in supplementary table S2. BTNX kit 2 and the SD assay demonstrated 100% (97-100%) specificity.

### 3.4. IgM detection

IgM detection was not evaluated in this study but results were recorded for BTNX, ACON and SD assays and are listed in supplementary table S1.

## 4. Discussion

As many manufacturers apply for EUA for COVID-19 serology assays from the FDA, it is becoming apparent that assay performance characteristics are variable (Tang et al., 2020, Theel E. S. et al., 2020). At the time of writing, 24 EUAs had been issued for serology tests; of these, six are LFIAs (RightSign COVID-19 IgG/IgM Rapid Test Cassette, Hangzhou Biotest Biotech Co., Ltd.; LYHER Novel Coronavirus (2019-nCoV) IgM/IgG Antibody Combo Test Kit (Colloidal Gold), Hangzhou Biotest Biotech Co., Ltd.; COVID-19 IgG/IgM Rapid Test Cassette, Healgen Scientific LLC.; Anti-SARS-CoV-2 Rapid Test, Autobio Diagnostics Co. Ltd.; Biohit SARS-CoV-2 IgM/IgG Antibody Test Kit, Biohit Healthcare (Hefei); and qSARS-CoV-2 IgG/IgM Rapid Test, Cellex Inc.)(FDA, 2020). Furthermore, although a number of LFIAs have been evaluated in the literature, some have been used in the context of seroprevalence studies without available peer reviewed data on their performance. Clinical evaluations of serologic assays for SARS-CoV-2 have been primarily focused on automated assays. Here we evaluated four LFIAs for their capacity to detect anti-SARS-CoV-2 IgG in retrospective serum, plasma, and whole blood specimens, focusing on sensitivity at ≥14 days after symptom onset in an inpatient cohort and on specificity.

The sensitivity of the LFIAs was evaluated against the Abbott SARS-CoV-2 IgG EUA assay, for which several peer reviewed studies have reported acceptable performance (Bryan et al., 2020, Tang et al., 2020, Theel Elitza S. et al., 2020). Although eight “BTNX kit 1” specimens were false negatives; all but one were collected <14 days after symptom onset. Further, four of them were collected from a single patient between one and two days post symptom onset; another specimen collected from this same patient two days after symptom onset generated a positive result. Similarly, for the remaining four false negative specimens (three patients), an additional specimen drawn from the same patients on the same day tested positive. Although it is surprising that an antibody response should be seen at all as early as one day into the disease course, it should be noted that this was a predominantly hospitalized and older cohort (Bryan et al., 2020), which may account for potentially incomplete clinical histories in some cases. Also, it has been documented that the infection may be asymptomatic for 1-2 weeks, so an immune response may be well underway by the time of symptom onset. Some specimens from patients who were PCR positive were negative by all assays, suggesting they had not yet generated detectable levels of antibodies to the viral antigens in the kits. It may take up to 21 days or more for some patients to develop a detectable antibody response (Yongchen et al., 2020).

In some cases, the LFIA tests detected a positive result sooner in a serial sampling series than the Abbott test did. Though these represent clinically positive results, against the Abbott assay, they would be “false positives.” Of the samples generating “false positive” results with BTNX kit 1, three were collected from a single patient one and two days post-onset of symptoms and with a PCR Ct of 29 (Panther Fusion® SARS-CoV-2 Assay); this patient went on to seroconvert on the Abbott assay 10 days later. Another of the “false positive” specimens, collected 10 days post-onset of symptoms, was associated with an Abbott OD value of 0.96 (manufacturer cut-off is 1.4); however, a specimen collected the following day from this patient was positive on the Abbott assay (OD, 2.33). A recent study has suggested that it may be beneficial to report OD ratios of 0.8-1.5 on this assay as inconclusive with a recommendation for repeat testing (Bryan et al., 2020). The remaining “false positive” specimen was collected on the day of symptom onset and a specimen collected 4 days later was positive on the Abbott assay. These early “false positive” results may be the result of reactivity with low-avidity IgG, that is not detected by Abbott.

Our approach to efficiently evaluate specificity was to test a set of pre-pandemic stored samples and then target a set of samples that contain potentially cross-reactive substances based on seasonal CoV or common interfering substances for serologic assays. The resulting specificity data from our sample set is promising, though somewhat limited in number. In particular, for three of the four assays tested, we did not observe any false positive IgG results from specimens from patients with a known history of seasonal coronavirus infection; these antibodies have been detected in a high proportion of individuals aged over 50 (Gorse et al., 2010) and cross-reactivity has been reported with other assays (Demey et al., 2020). One specimen from a patient with a recent history of coronavirus HKU1 infection tested positive on the ACON assay; however, 12 additional specimens from individuals with a history of HKU1 infection did not cross-react in this assay. We did observe false IgM positive results with all assays where this was tested, but given the questionable clinical significance of IgM detection (Farnsworth and Anderson, 2020, Landry, 2016), the propensity for both antibodies to become detectable within a similar time frame, and the variability over the serial samples included in our data set, we opted to evaluate the performance of IgG detection in these assays only. False positivity due to autoantibodies has been reported for some SARS-CoV-2 serology assays (Theel E. S. et al., 2020); in our study, samples positive for rheumatoid factor generated positive IgM results on both assays, but IgG remained negative.

One potential use for these assays might be to confirm antibody production in patients with resolved symptoms, independent of disease detection by a SARS-CoV-2 PCR assay or not. In this case, after a 14 day self-quarantine, would these serological tests detect a positive result? The results of this study of sera from a primarily hospitalized population show that the sensitivity of IgG detection at 14 days or more post-symptom onset was 95% in two cases (BTNX kit 1, and ACON). When compared directly with Abbott results, sensitivity increased to 99% for both of these assays; similarly, BTNX kit 2, and SD sensitivity was 94% and 100%, respectively. These LFIA tests show good performance for a 15-minute test that is very easy to perform; however, BTNX kit 1 and ACON were the only assays to generate false positive IgG results, supporting the theory that assays providing higher sensitivity may come with a compromise in specificity. Nonetheless three of the kits tested did not show any false positives in a sample set that included a diverse representation of potential cross-reactivity, and specificity for any assay did not fall below 98%. Additional studies will be needed to determine if this measure of sensitivity holds true for more mild disease, and whether sensitivity may increase (or decrease) past 14 days. Other studies have shown improved sensitivity after 17 days from symptom onset (Bryan et al., 2020). While no serologic test is perfect, these results are encouraging that rapid and simple tests can provide an adequate level of sensitivity and specificity. Importantly, several other LFIAs tested by our group showed poor sensitivity and/or specificity (data not shown), indicating the importance of rigorous validation prior to implementation in any setting. It is likely that all tests will have a measurable false-positivity rate, but our results suggest that a substantial number of samples from patients with a history of seasonal CoV or even other viral infections will be required to better define the rate of false-positivity. Even if some tests maintain a high (>99%) specificity, the individual patient may be best served by an orthogonal approach to testing, whereby two methods that target different antigens (whether two LFIAs or an LFIA followed by an EIA or CIA) are used to increase positive predictive value for predicting true exposure to SARS-CoV-2. However, manufacturers are only obliged to disclose the nature of their assay target(s) upon EUA issuance, so the role of the many pending assays in this approach is currently unclear. The sensitivity of the LFIAs characterized herein suggests that such an approach would have only a minor impact on clinical sensitivity overall by using two assays. This concept is supported by the fact that of the false negatives, four samples were not detected by any of the assays, demonstrating that most positive samples were detected by all assays.

One strength of the study is that among the specificity sample set, we included 27 samples from patients who had recently experienced seasonal CoV. While additional studies are required to focus on other patient groups and sample types, the sensitivity sample set in this study was already larger than the data listed for 10 of 13 EUA approved assays and the specificity samples set was similar to 4 of 13 EUA approved assays, current as of June 8, 2020 (FDA, 2020). The primary weaknesses include the positive samples from a primarily hospitalized cohort, the retrospective nature of samples (including freeze/thaw), and the lack of fingerstick blood samples, for which many of these assays are designed. In a pandemic, reliable information is essential to public health responses and individual healthcare decisions. These results suggest that, with further investigation/data/study/evaluation, LFIAs could potentially be used to meet that need, particularly in low-resource settings or those with limited access to health care. Importantly, it must be noted that detection of IgG does not mean that neutralizing antibodies are present. There is not yet sufficient data in the literature to determine whether detection of IgG may (or may not) correlate with immunity or protection of future exposure to SARS-CoV-2.

## Data Availability

There are no external datasets associated with this manuscript. All data can be found in the manuscript and supplementary material.

## Supplementary File

Table S1. SARS-CoV-2 IgG and IgM results for the lateral flow immunoassays evaluated in this study

Table S2. SARS-CoV-2 false positive IgG results detected during this evaluation Figure S1. An invalid result observed during this evaluation

## Acknowledgements

We would like to thank the Mayo Clinic Center for Individualized Medicine for their continued support and the Mayo Clinic Arizona Department of Laboratory Medicine and Pathology staff for all their hard work supporting this study and the care of our patients during this pandemic. We would also like to thank the manufacturers for supplying some of the kits (ACON and BTNX kit 1). We also thank Safe Health Systems who supplied some kits (SD and BTNX kit 2) as part of a joint partnership with Mayo Clinic.

